# Neurocognitive measures of self-blame and risk prediction models of recurrence in major depressive disorder

**DOI:** 10.1101/2021.01.13.21249739

**Authors:** Andrew J. Lawrence, Daniel Stahl, Suqian Duan, Diede Fennema, Tanja Jaeckle, Allan H. Young, Paola Dazzan, Jorge Moll, Roland Zahn

**Author notes:** Corresponding author Dr Roland Zahn (see address above).

## Abstract

**Background:** Overgeneralised self-blaming emotions, such as self-disgust, are core symptoms of major depressive disorder (MDD) and prompt specific actions (i.e. “action tendencies”), which are more functionally relevant than the emotions themselves. We have recently shown, using a novel cognitive task, that when feeling self-blaming emotions, maladaptive action tendencies (feeling like “hiding” and like “creating a distance from oneself”) and an overgeneralised perception of control are characteristic of MDD, even after remission of symptoms. Here, we probed the potential of this cognitive signature, and its combination with previously employed fMRI measures, to predict individual recurrence risk. For this purpose, we developed a user-friendly hybrid machine-/statistical-learning tool which we make freely available.

**Methods:** 52 medication-free remitted MDD patients, who had completed the Action Tendencies Task and our self-blame fMRI task at baseline, were followed up clinically over 14-months to determine recurrence. Prospective prediction models included baseline maladaptive self-blame-related action tendencies and anterior temporal fMRI connectivity patterns across a set of fronto-limbic a priori regions of interest, as well as established clinical and standard psychological predictors. Prediction models used elastic-net regularised logistic regression with nested 10-fold cross-validation.

**Results:** Cross-validated discrimination was highly promising (AuC≥0.86), and positive predictive values over 80% were achieved when including fMRI in multi-modal models, but only up to 71% (AuC≤.74) when solely relying on cognitive and clinical measures.

**Conclusions:** This shows the high potential of multi-modal signatures of self-blaming biases to predict recurrence risk at an individual level, and calls for external validation in an independent sample.

## Introduction

One central feature of cognitive models of vulnerability to major depressive disorder (MDD) is a tendency to excessively blame oneself (1). These models have pointed to over-generalised self-blame as causing worthlessness and hopelessness, core symptoms of MDD that are distinctive compared with other recurrent emotional disorders such as panic disorder (2). Self-blame in MDD is strongly associated with complex emotions such as guilt, self-disgust, and shame(3). Social psychologists have highlighted that specific maladaptive “action tendencies” such as “feeling like hiding” were associated with self-blaming emotions(4) and distinguish adaptive from depressogenic self-blame more accurately than the label of the emotion(5). Indeed, using a novel Action Tendencies Task, we have shown that self-blame-related action tendencies, in particular feeling like creating a distance from oneself and hiding, as well as an overgeneralised perception of control for other people’s wrongdoing were distinctive of remitted MDD patients compared with control participants cross-sectionally (6). If measures of maladaptive self-blame-related action tendencies prospectively predicted future recurrence risk in remitted MDD, this would provide critical evidence for their role in MDD vulnerability. Apart from its pathophysiological importance, the identification of novel predictors of recurrence is needed for developing accurate risk prediction tools, because clinical variables are poor predictors at an individual level(7) and there are no established biomarkers(8).

Over the last decade, the application of machine learning to MRI-based prediction of individual responses to antidepressant treatments(9-12) has raised great promise for developing predictive markers in MDD. Machine learning models are powerful at making personalised predictions, because they condense multiple variables, such as MRI signal in different brain regions, or different modalities (e.g. cognitive, structural MRI & functional MRI(10)) into one model used to classify individuals based on complex interactions between the multiple input variables(13). This approach has been used to predict whose symptoms will respond to a treatment (9, 10, 14, 15). Successful development of novel treatments and stratification algorithms to improve long-term outcomes of recurrent MDD, however, requires predictors that capture the neurocognitive underpinnings of asymptomatic precursors of recurrent symptoms. Using a standard general linear model, we have previously identified an fMRI signature of MDD patients who will develop a recurring episode after recovering from previous depression over one year of follow-up(16), but the machine learning approach we used to show its predictive value at the individual level with a positive predictive value of 74% had several weaknesses: Firstly, it used a high variance leave-one-out cross-validation method(17, 18). Secondly, it used the extracted clusters showing the most significant association with recurrence risk in our SPM general linear model. This overestimates model performance due to selecting voxels and variables on the basis of another model comparing recurring episode and stable remission patients in the same sample(19). Here, we therefore sought to overcome these weaknesses.

As recently reviewed(20), guilt-proneness has been previously selectively associated with subgenual cingulate cortex (SCC) activation on fMRI(21), whereas frontopolar activation was most reproducible for guilt vs. other-blaming emotions irrespective of individual differences in guilt-proneness(22-24). The latter is part of the “Default-Mode Network” and its connectivity with the SCC has been implied in depressive rumination (25), which is typically of a self-critical nature. Interestingly, the SCC was abnormally active in current MDD and normalisation of its metabolism was associated with remission(26, 27), its connectivity predicted response to psychological vs. antidepressant treatment (14) suggesting its central importance. The SCC, however, is only part of a network of regions involved in self-blaming feelings. Using fMRI, we have demonstrated that proneness to self-blaming feelings in healthy people was associated with increased functional connectivity between the SCC and the right superior anterior temporal lobe (ATL(28)), which we had previously demonstrated to enable differentiated interpretations of the meaning of social behaviour(29, 30) (e.g. differentiating actions as “impolite”, or “absent-minded”, rather than just over-generally “bad”).

In an independent previous sample of patients with remitted MDD, we demonstrated abnormal functional connectivity between the right superior ATL and fronto-limbic networks when experiencing self-blaming feelings relative to feelings related to blaming others (compared with never-depressed controls, independently of medication status(24)). These fronto-limbic networks included the SCC, frontopolar cortex, hypothalamus, and hippocampus. This concorded with Price & Drevets’ neuroanatomical model of mood disorders which includes the superior ATL(26) because of its close anatomical connection with the medial prefrontal network including the subgenual cortex and the hypothalamus, thereby providing a crucial and often forgotten link between the limbic forebrain (including the hippocampus) and frontal cortex. The ATL is probably under-reported, because fMRI in this region requires optimised imaging parameters due to the abundance of artefacts(31).It is notable that the original formulations of the “Default Mode Network” based on positron emission tomography included the superior ATL(32). Our focus on the right superior ATL is based on its predominance for socio-emotional functions(30, 33) (22, 30, 34-36) relative to the left ATL. This concords with the efficacy of right temporal electroconvulsive therapy in MDD(37), response to which was predicted by right ATL-frontal connectivity including the SCC(38).

The aim of the current study was to develop a user-friendly prediction tool and use it to provide robust estimates for the potential to predict MDD recurrence risk at the individual level, when using cognitive and fMRI-based signatures of self-blaming biases. Further, as one could argue that fMRI is relatively expensive for widespread use, we also probed the potential to replace it by our novel cognitive task for future clinical decision support systems.

## Methods and Materials

### Participants and standard measures

Participants were recruited as a part of a prospective cohort study (from 2011-2014) in a clinical research facility ((16), ethics reference: 07/H1003/194). All participants provided informed consent (verbal for initial telephone screening and written otherwise). Recruitment has been described previously (39). Inclusion criteria were: MDD according to DSM-IV-TR(40), remitted for ≥six months, psychotropic medication-free, right-handed, native English speaking, with normal or corrected-to-normal vision. Main exclusion criteria were: current Axis-I disorders including a history of substance or alcohol abuse, and past comorbid Axis-I disorders being the likely cause of depressive symptoms. Patients were subsequently followed up clinically over 14 months using the well validated Longitudinal Interval Follow-up Evaluation interview for DSM-IV (LIFE-IV(41)). Raters were blinded to the baseline results; inter-rater reliability was excellent (16).

For our multi-modal prediction model we used a complete-cases analysis (see supplemental methods) including n=52 MDD with complete follow-up data that we were able to categorise into recurring episode over 14 months (n=18) vs. stable remission with no recurring episode (n=34).

The following standard clinical and psychological measures were selected for use in the prediction model as based on previous reports establishing them as reproducible predictors of MDD recurrence at the group level: number of previous episodes ((42), categorised into non-recurrent [i.e. 1 previous episode], recurrent [2-4 episodes], and highly recurrent [>5 episodes]); Beck Depression Inventory (BDI, (43)) as a measure of residual symptoms which are associated with recurrence risk (42) and which loads heavily on self-criticism, also associated with recurrence (44); Global Assessment of Functioning to capture co-morbidity and functional impact (45); Rosenberg Self-Esteem (46) (47); and Positive and Negative Affect Schedule (48), where the Negative Affect subscale was used as a measure of negative emotionality, closely related to “neuroticism” (49) and the Positive Affect subscale used because of its negative association with subsequent recurrence(50) in previous studies.

### Experimental Stimuli

Stimuli for both fMRI and behavioural tasks were written sentences each presenting an abstract hypothetical social behaviour contrary to social and moral values. Participants were asked to imagine the situation in each stimulus for two conditions differing by the agency of the participant. In the self-agency condition the participant was described to act towards their friend, in the other-agency condition their friend acted towards them. Stimuli were based on the value-related moral sentiment task (VMST) used previously (16, 39). The same social behaviours were used in both conditions (90 trials per condition, 50% *per se* negative social behaviours [e.g. does act stingily] and 50% negated positive behaviours [e.g. does not act generously]). Participants were asked to provide the name of their best friend of the same gender, with whom they were not related and not romantically involved.

After the fMRI and behavioural tasks, participants were asked to rate all items in the self- and other-agency conditions for unpleasantness (“How strongly would you feel unpleasant feelings?”: scale of 1 [“Not unpleasant”] - 7 [“Extremely unpleasant”]). As in our previous paper (16), we restricted our analyses to the items deemed most emotionally relevant to each participant (defined as items rated equal or higher than the individual’s median unpleasantness rating for each condition: self- and other-agency).

### fMRI acquisition and paradigm

As previously described (16), we used an fMRI protocol optimised for detection of ventral brain regions (see Supplemental Methods) and standard fMRI results were previously reported (16). Participants were presented with hypothetical social actions in self-agency and other agency conditions (stimuli described above). Stimuli were presented for 5 seconds in three runs in pseudorandom order, runs were counterbalanced across participants, and interspersed with a baseline visual fixation of pattern condition (n=90). In the scanner participants were asked to decide whether each situation would feel mildly or very unpleasant to ensure they paid attention to the task and that they make an emotional decision about the stimuli. We used a jittered (range 500ms) inter-trial interval with mean duration four seconds.

### fMRI analysis

As in our previous analysis [(16), Supplemental Methods] to measure functional connectivity, we employed the well-established Psychophysiological interaction (PPI) analysis (51), which requires the extraction of the right superior ATL signal time course (physiological variable, as previously: (28); 6mm radius sphere; MNI coordinates: x=58, y=0, z=-12) and the creation of an interaction term with the psychological variable (the contrast between the most highly unpleasant items in the self-agency condition vs. the visual fixation condition and highly unpleasant items in the other-agency condition vs. the visual fixation condition).

Regression coefficient averages over a priori ROIs (depicted in Figure 1b and further described below) as defined previously using an independent sample(24) were extracted as raw values using MarsBaR (52) and used to capture self-blame-related fMRI connectivity with our ATL seed region, as well as ATL connectivity with these ROIs irrespective of psychological condition, whilst co-varying for root mean square movement parameters during the scan (obtained from the realignment process). In addition, we modelled standard Blood Oxygenation Level Dependent (BOLD) effects following the modelling approach as previously described (16) for self-blame in the SCC(27) and ATL seed region. This was in keeping with the PPI approach(51). To limit the number of variables, we modelled BOLD only in our primary regions of interest (SCC and ATL).

**Figure 1.**
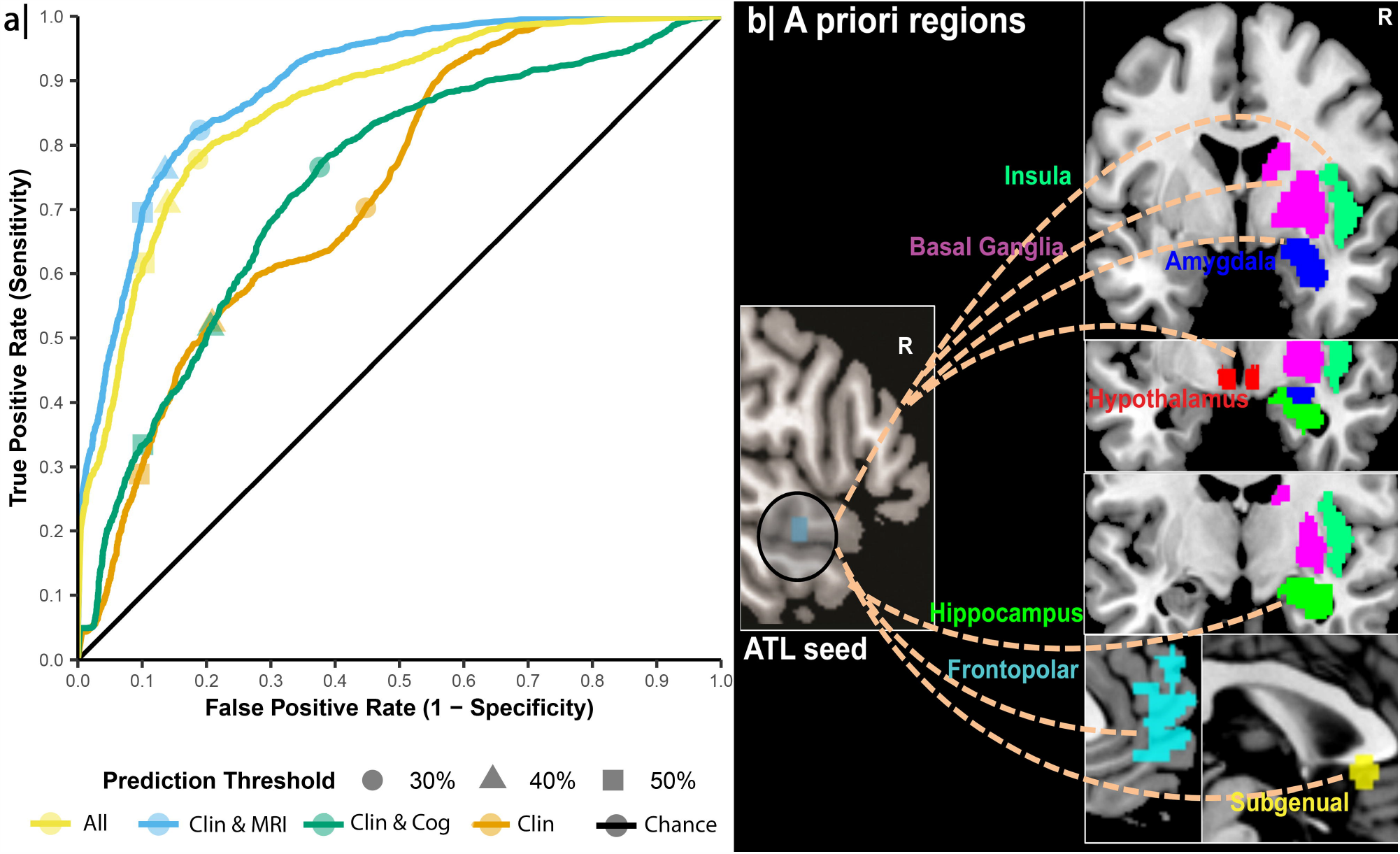
**a** Average cross-validated Receiver Operator Characteristic (ROC) curves for repeated nested regularised logistic regression models (the same n=52 MDD participants in all models, 30 nested inner and 100 outer loop repetitions using 10-fold cross-validation, R-software).The *clinical* variables model included standard scales and known clinical predictors. *Cognitive & Clinical*: maladaptive self-blaming action tendencies measures were added. *fMRI & Clinical*: fMRI measures were added. *All modalities*: included Cognitive, Clinical and fMRI measures. **b**| All a priori ROIs reported in a previous independent paper (24) with close anatomical connections to our ATL seed region and of sufficient anatomical specificity for data extraction were included (i.e. regions spanning multiple Brodmann areas [BA]such as the dorsolateral frontal cortex were discarded). The model included midline subgenual cingulate cortex (SCC), frontopolar cortex (BA10), and hypothalamus, as well as right hemispheric hippocampus, amygdala, insula, and basal ganglia (striatum/pallidum) ROIs to match our a priori right hemisphere ATL seed region (24).

### fMRI regions of interest

To ensure independence of our ROIs from the current dataset, we used all a priori ROIs from our previous independent publication (24), but in order to reduce the number of ROIs, we chose only right sided ROIs reflecting our right sided ATL seed region, except for the midline regions, where we used bilateral ROIs (see Supplemental Methods).

### The Action Tendencies Task

As previously described (6), for each described social behaviour, the participant was asked: “What would you feel like doing in response to this behaviour?”. “Please select the option that you feel that you would most strongly feel like doing (see Supplemental Figure 1): “verbally or physically attacking/punishing your best friend”, “verbally or physically attacking/punishing yourself”, “apologising/fixing what you have done”, “hiding”, “creating a distance from your best friend”, “creating a distance from yourself”, “no action”, “other action”. They then rated the hypothetical situation regarding how much control they feel they would have (1:None at all → 7:Complete).

We have previously reported that self-agency-related hiding and self-distancing, as well as an overgeneralised perception of control for others’ wrongdoing was characteristic of MDD at baseline compared with control participants (6). In the present paper, we report their association with recurrence risk for the first time. As previously (6), we separately calculated the proportion of trials that “hiding” or “self-distancing” were selected in the self-agency condition for the highly unpleasant items as defined using individual median splits as described above. In order to reduce the number of variables, we z-transformed these proportions across the whole sample to combine hiding and self-distancing into a single average of z-scores of maladaptive self-blaming action tendencies. We measured over-generalised perception of control as previously by subtracting each subject’s average perceived control rating in the highly unpleasant other-agency condition trials from the same quantity in their highly unpleasant self-agency trials. Perceived control is expected to be higher during self-agency, so a smaller value in the calculated difference measure indicates abnormally higher perception of control in the other-agency condition which is conceptually related to the notion of “Omnipotent Responsibility Guilt” (i.e. caused by an exaggerated sense of responsibility for others wellbeing) as shown to be associated with MDD (53).

### Prediction modelling

We developed a software tool (AL) to implement the proposed methods and make this tool available for use by the scientific and clinical community as an R-package (https://www.github.com/AndrewLawrence/dCVnet). We adopted a hybrid approach combining statistical- and machine-learning to balance their benefits and limitations in this type of prediction problem. In contrast to traditional statistical methods we reduced overfitting and allowed correlated predictors through regularisation. We also carefully cross-validated performance by including all modelling decisions within the cross-validation. This hybrid approach can likewise be contrasted to more complex (and data-hungry(54)) machine learning methods: each model uses relatively few of the available predictors which are chosen a-priori based on previously published results and clinical utility. To further reduce complexity, we used a model which assumes the effects of the predictors are linear and additive, any interaction terms or predictor transformations must be prespecified. The software tool is called dCVnet, for “<u>d</u>ouble <u>C</u>ross-<u>V</u>alidation for the elastic-<u>net</u>”. It employs elastic-net regularised logistic regression with double (also termed nested) cross-validation, consisting of an ‘outer’ cross-validation of model performance measures with an ‘inner’ cross-validation to independently tune elastic net hyperparameters.

#### Elastic-net model

Elastic-net regularised binary logistic regression extends logistic regression with regularisation. This acts as a penalty to model complexity and reduces this by shrinking coefficients towards zero(55). The elastic-net regularisation penalty comprises two types of penalty with different roles: the L2 (Ridge) penalty produces allows correlated predictors to jointly enter the model and stabilises solutions; while the L1 (LASSO) penalty encourages variable selection (56). The amount of regularisation (hyperparameter: lambda) and balance of the two types of regularisation penalty (hyperparameter: alpha, indicating the fraction of the penalty that is of LASSO-type) are hyperparameters of the elastic-net procedure in that they determine the learning performance of the algorithm and tuning alpha and lambda adapt the algorithm to different problem settings.

#### Cross-validation & Tuning

Prediction models tune to both the reproducible relationships between data and outcome (desirable) and the idiosyncrasies of the particular dataset they are trained on (undesirable). The latter is worse with small data and complex/flexible models and results in overfitting - a high generalisation error when the model is provided new, data to predict for. Although external validation is preferable, internal validation, involving (re-)training the model on a subset of the available dataset and validation of performance in the remainder, is often the only practical option and still allows nearly unbiased estimates of the ‘true’ or generalisation error (17). Overfitting does not just affect the identification of model coefficients, overfitting occurs for model selection and hyperparameter tuning(57). For cross-validation to combat this inflation of predictive performance model selection and hyperparameter tuning must be conducted independently within the internal validation(57). We implemented such a procedure in dCVnet. Specifically, double (also called nested) cross-validation is used to obtain performance estimates without optimistic bias at the same time as tuning hyperparameters based on cross-validated performance(57). For both inner and outer cross-validation loops our tool employed repeated k-fold cross-validation, an unbiased estimate of expected (out-of-sample) prediction error with lower variance than bootstrap, hold-out, and leave-one-out cross-validation methods(56, 58). For this particular application of dCVnet, stable hyperparameter selection could be obtained (for all models) with 30 repetitions of 10-fold cross-validation, and stable cross-validation results with 100 repetitions of 10-fold cross-validation. For tuning, six logarithmically spaced values of alpha were considered between 0.01 (mostly Ridge) and 1.0 (a LASSO model). For each alpha, one hundred lambda values were determined from the data by glmnet(59). The tuned alpha and lambda were selected based on the minimum mean square error (MSE; also termed Brier score), rather than AuC, as MSE combines discrimination and calibration components of prediction performance.

#### Software

dCVnet was programmed in R-software version 3.6.2 and makes repeated calls to the glmnet 3.0.2 package’s binomial elastic-net fitting function to produce the multiple models required for nested k-fold cross-validation(59). The novelty of dCVnet lies in providing a documented and standardised implementation of this particular machine learning pipeline making it accessible to researchers lacking programming experience required for more general machine-learning software environments (e.g. sklearn, caret, tidymodels or mlr). We make this software including documentation available (<u>github.com/andrewlawrence/dCVnet</u>) to improve reproducibility and future open source development of clinical decision support systems.

#### Model evaluation

Predicted probabilities of depression recurrence were compared with the known recurrence status to calculate a variety of cross-validated prediction performance measures(60). These including common classification metrics (such as Positive Predictive Value, PPV; Negative Predictive Value, NPV) based on a 50% classification cut-off. For PPV and NPV we adjusted our estimates for the 42% prevalence of recurrence in the wider study sample. We further considered threshold-independent performance measures: the Brier score (mean squared difference between predicted probability and binary outcome), and the Area under the receiver-operator characteristic Curve (AuC; also termed the concordance statistic). Finally, we considered model calibration (i.e the fidelity of the predicted probability to the observed proportion with the outcome) by calculating the intercept and slope of the calibration graph(60). Consistent with our aims, at this point in model development the measure of prime interest is the AuC as this reflects the discriminative potential of a model independent of the classification threshold, or model calibration.

## Results

As shown in Table 1, there were no demographic difference between groups. As expected, the recurring episode group showed a higher BDI score at baseline. We obtained an AuC of at least .86, ≥80% cross-validated accuracy and moderate calibration for classifying patients into Recurring Episode vs. Stable Remission when combining multi-modal information from fMRI & standard clinical and psychological measures, with or without our novel Action Tendencies Task (Table 2, Figure 1). We further probed the contributions of the different modalities to predictive performance. Relying solely on standard clinical and psychological measures achieved an AuC of .73 and positive predictive values of 68% (Table 2). Our results showed, however, that the novel cognitive measure, our Action Tendencies Task, did not improve prediction performance to a relevant degree (AuC=.74, positive predictive values of 71%, Table 2). We therefore also ran a prediction model which only contained the Action Tendencies Task variables to demonstrate that they were associated with recurrence risk (AuC=.67, Table 2), but this model performed slightly worse compared with known clinical and standard measures.

**Table 1.** Demographic characteristics and symptoms

|  | Remitted MDD |  | p-overall |
| --- | --- | --- | --- |
|  | Recurring<br>n=18 | Stable<br>n=34 |  |
| Age | 35.9 (13.2) | 34.9 (13.9) | 0.79 |
| Sex |  |  | 0.98 |
| Female | 13 (72.2%) | 23 (67.6%) |  |
| Male | 5 (27.8%) | 11 (32.4%) |  |
| Years of Education | 16.1 (2.08) | 17.1 (1.88) | 0.11 |
| BDI Score | 5.44 (3.81) | 2.97 (3.11) | 0.03* |
| MADRS Score | 1.22 (1.56) | 1.03 (1.49) | 0.67 |
| GAF Score | 83.6 (6.46) | 86.5 (4.84) | 0.10 |

MDD = major depressive disorder; BDI = Beck Depression Inventory; MADRS = Montgomery-Asberg Depression Rating Scale; GAF - Global Assessment of Function. Standard deviations are displayed in parentheses. 52 fully remitted MDD patients had available baseline data for all predictors included in the multi-modal prediction model and were followed up clinically over 14 months to determine recurrence. As expected, there were slightly higher residual depressive symptoms on the more sensitive BDI in the group with a subsequent recurrence.

**Table 2.** Cross-validated Prediction Model Results

| Metric | Cognitive | Clinical | Clinical &<br>Cognitive | Clinical &<br>fMRI | All modalities |
| --- | --- | --- | --- | --- | --- |
| Accuracy | 0.68 | 0.69 | 0.70 | 0.83 | 0.80 |
| Sensitivity | 0.33 | 0.28 | 0.33 | 0.69 | 0.61 |
| Specificity | 0.86 | 0.91 | 0.90 | 0.90 | 0.90 |
| PPV | 0.63 | 0.68 | 0.71 | 0.83 | 0.81 |
| NPV | 0.64 | 0.63 | 0.65 | 0.80 | 0.76 |
| AuC | 0.67 | 0.73 | 0.74 | 0.90 | 0.86 |
| Brier Score | 0.20 | 0.20 | 0.20 | 0.13 | 0.15 |
| Calibration Intercept | -0.12 | -0.02 | -0.10 | 0.02 | -0.08 |
| Calibration Slope | 0.78 | 0.99 | 0.83 | 0.61 | 0.55 |

Cross-validated (100x repeated 10-fold CV) performance measures from the different prediction models. PPV=positive predictive value, NPV=negative predictive value, both were adjusted to the 42% prevalence found in the full sample (16). AuC=area under the Receiver Operating Characteristic curve. Cognitive=Self-blame-related Action Tendencies Task variables, Clinical=Standard psychological and clinical measures, fMRI=self-blame-related fMRI variables.

When comparing unregularised odds ratios from single-predictor logistic regressions with the regularised odds ratios, there was relatively little shrinkage of coefficients (i.e. a small lambda was chosen), reflecting the relatively good cross-validated performance of the model (Table 3). Interestingly, there were predictors which on their own had no relevant association with recurrence risk, but contributed to the multivariate prediction. Maladaptive action tendencies were associated with recurrence risk by themselves and in the full regularised model (Table 3), whereas overgeneralised perception of control was not (Table 3).

**Table 3.**
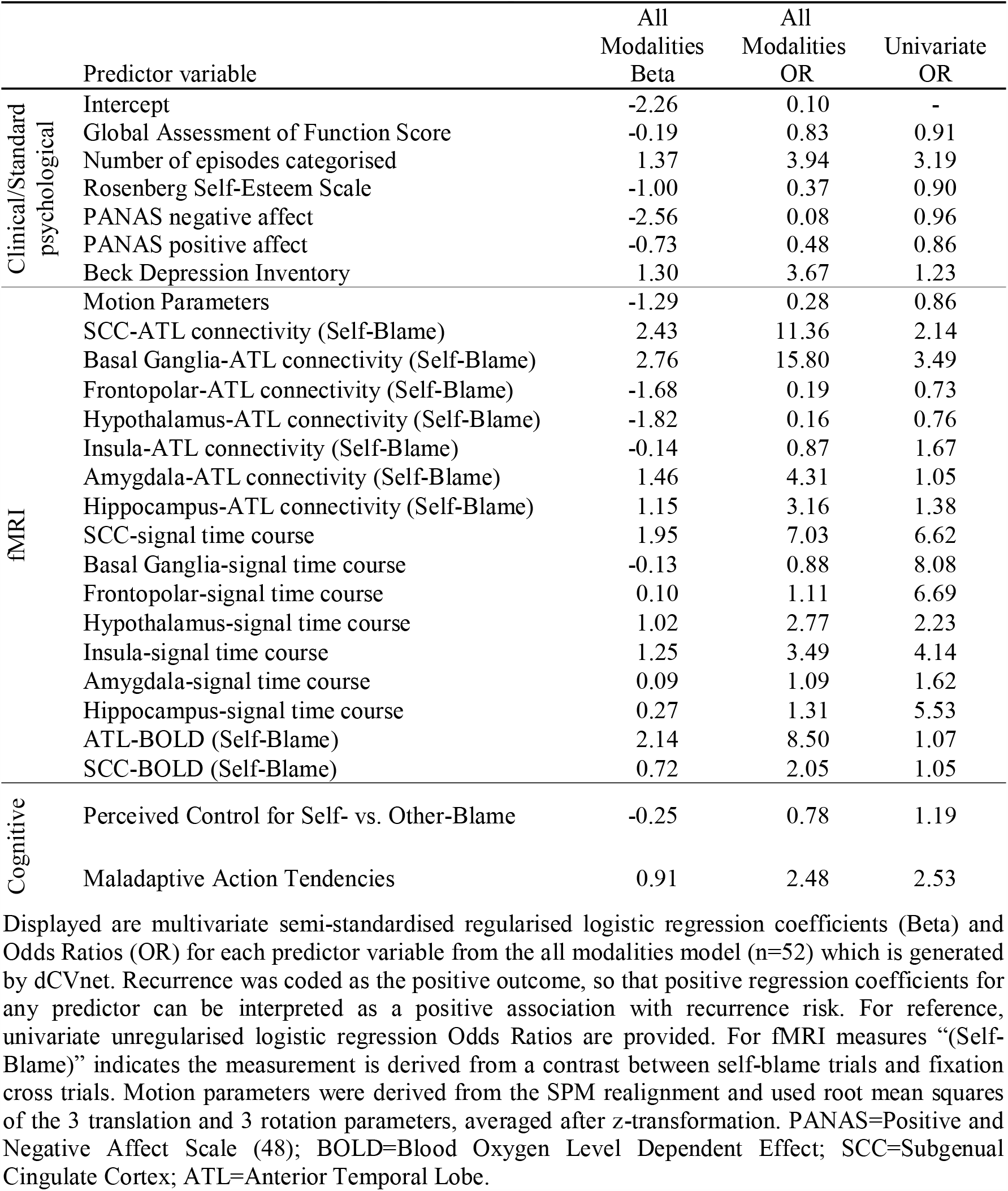
Model coefficients predicting recurrence

## Discussion

We confirmed that combinations of clinical, psychological and self-blame fMRI measures were highly promising candidates for predicting individual recurrence risk in MDD. Using a principled hybrid statistical-/machine-learning approach we observed excellent cross-validated discriminative performance with an AuC of 0.9 and positive predictive values above the suggested benchmark of 80% for clinically useful markers (61). In contrast, our novel cognitive task to capture self-blame-related action tendencies was not sufficient to replace fMRI. We nevertheless obtained AuC values above .7, which is considered “fair” discriminatory value (62) and there was an association of maladaptive action tendencies with subsequent recurrence risk (odds ratio above 2.5). Yet, there was no relevant improvement on known clinical and standard measures regarding its discriminative value. These findings of modest predictive utility are in keeping with the previous literature showing that clinical and standard measures in themselves are unable to make accurate predictions of individual recurrence risk (7, 63).

Despite the lack of evidence for the predictive utility of the Action Tendencies task in this dataset, there are several strategies which could be pursued to improve the value of experimental cognitive tasks in future prediction models. Firstly, our task was purely text-based and lacked immersive features. Immersion could be improved by adopting a virtual reality paradigm (64). Secondly, any single task is unlikely to capture all relevant aspects of vulnerability and thus future work could integrate a wider range of cognitive measures, for example a recently developed cognitive task measuring effort and reward decisions has shown promising but modest effect sizes for predicting recurrence in MDD (65) resulting in similar AuC values of around .70 as our action tendencies task, its combination with action tendencies may out-perform the separate models.

Although a wide variety of clinical and standard measures with known predictive value were assessed, performance of the model using just these features was modest (AuC = 0.73) and one might ask whether important predictors were omitted. Recently, promising results were shown for recurrence risk prediction using a machine-learning model which included childhood trauma(66, 67), although this needs further replication, we did not measure childhood trauma and adding this to our prediction model may improve its accuracy in the future. Despite their theoretical importance (2), our model did not include stressful life events during the follow-up period, because our aim was prospective prediction from baseline data. We note that although 60% of our 93 patients with completed follow-up data in the overall study reported stressful life events during the 14-month follow-up period (as determined by clinical interview and standardised questionnaires), there was no difference in the rate of occurrence of these events between the *Recurring Episode* (61%) and *Stable Remission* (59%) groups. This is consistent with the hypothesis that life events trigger recurrence not directly, rather through their interaction with other factors, such as self-blaming biases(2), a hypothesis which merits further investigation.

Previous studies using machine learning approaches to imaging data in depression, have usually relied on voxel-based methods (68). Yet, these methods do not allow integrating cognitive and clinical variables in a straightforward way. This is why we decided to employ a simpler approach to the imaging analysis which relies on ROIs and this may be more feasible for future clinical applications. By employing the elastic net as a machine learning extension of logistic regression, we alleviated overfitting by shrinking the regression coefficients towards zero, this allows for automatic variable selection by omitting some predictors. The nested cross-validation is vital to provide realistic estimates of out-of-sample prediction accuracy and thereby estimate “internal validity”(69), a shortcoming of many studies of clinical predictors which commonly employ shared cross-validation for both model tuning and performance estimation. Although this prognostic model is at an early stage, our approach follows the MRC Prognosis Research Strategy Partnership (69) guidelines for developing and reporting prognostic models.

On a more cautionary note, for prognostic model development, our sample size was relatively small (70). Although we deliberately adopted more stable hybrid methods than the voxel-wise SVM considered by(70), we must nevertheless replicate these findings in a larger independent sample before drawing clinical conclusions. Further, we included patients who were fully remitted from symptoms and had no relevant co-morbidity, thus generalisation to patients with partial remission and comorbidity will need to be investigated.

## Conclusions

Maladaptive self-blame-related action tendencies and fMRI measures predicted subsequent recurrence risk in our MDD sample. When including fMRI, our statistical learning-based prediction risk tool showed promising potential to predict recurrence risk at an individual level. This calls for external validation in an independent sample.

## Data Availability

The dataset used in this publication is available upon request, subject to a Data Access Agreement.

http://doi.org/doi:10.18742/RDM01-482

## Acknowledgements

This study was funded by an MRC Clinician Scientist Fellowship to RZ (G0902304). RZ and DS were partly and AJL fully funded by the National Institute for Health Research (NIHR) Biomedical Research Centre at South London and Maudsley NHS Foundation Trust and King’s College London and by a NARSAD Independent Investigator Grant (24715) from the Brain & Behavior Research Foundation. The views expressed are those of the authors and not necessarily those of the NHS, the NIHR, or the Department of Health. J.M. was supported by the LABS-D’Or Hospital Network, Rio de Janeiro, Brazil. J.A.G. This study was funded by the UK Medical Research Council (G0902304). D.F. was funded by the Medical Research Council Doctoral Training Partnership (Ref: 2064430). We are grateful to Dr Karen Lythe for collecting the primary data.

