## Supplementary Material for "Neurocognitive measures of self-blame and risk prediction models of recurrence in major depressive disorder"

*Corresponding author

**Short Title:** *Self-blame and depression risk prediction*

*Keywords:* self-blame; guilt; anger; action tendencies; fMRI; depression; recurrence; moral emotion

### Supplemental Methods

#### *Exclusion reasons*

Our exclusion reasons for potential participants after pre-screening have been previously described (1). For the complete case analysis approach employed here, our initial sample consisted of n=108 MDD participants of whom n=81 had complete fMRI data (n=81/108), n=76 of 108 had complete Action Tendencies Task data, and n=87 of 108 had complete clinical and standard psychological measures. All measures were complete in n=63 MDD participants, of whom n=52 could be categorised into stable remission (i.e. Psychiatric Status Rating Scale [PSR, (2)] < 4 and not requiring treatment) and recurring episode (i.e. PSR > 4). 11 participants could not be used for this binary outcome definition, because they showed major symptoms (i.e. PSR=4) or required treatment despite lower symptom severity levels (PSR=3) and were excluded as for the primary analysis of our previous paper(1) in which we showed in a secondary analysis that the subthreshold symptom group resembled more the stable remission than the recurring episode group.

#### *fMRI acquisition*

As previously described (1), we used an fMRI protocol optimised for detection of ventral brain regions. T2*-weighted echo-planar images (3 runs of 405 volumes with 5 dummy scans) were acquired on an MRI scanner (3T Achieva, Philips) with an 8-channel head coil, 3mm section thickness, ascending continuous acquisition parallel to the anterior to posterior commissural line, 35-40 slices depending on the participant's head, repetition time=2000 milliseconds, echo time=20.5 milliseconds, field of view=220 x 220 x 120mm, acquisition matrix=80 x 80 voxels, reconstructed voxel size=2.29 x 2.29 x 3mm, and sensitivity encoding factor=2, enabling dynamic stabilisation to correct for signal drift. T1-weighted 3-dimensional MRIs were acquired for co-registration (further details in (1)).

#### *fMRI analysis*

We used our previous first level models (1), functional images were realigned, unwarped, coregistered to the participant's T1-weighted images, and normalised to the SPM8 template (used to be consistent with our previous paper) using the transformation parameters for the T1-weighted image, before applying a smoothing kernel of 6mm full-width-half-maximum (http://www.fil.ion.ucl.ac.uk/spm/). The first-level models used all default options, but included time and dispersion derivatives and included a grey matter mask previously described to exclude voxels outside the brain (3).

#### *fMRI regions of interest*

We used our previous SCC (6 mm radius sphere around -4, 23, -5 and its right hemispheric mirror coordinate combined). As previously described (3, 4), ROIs had been created using the Automatic Anatomical Labelling atlas (AAL (5)) implemented in the Wake Forrest University (WFU) Pickatlas tool ((6), for details see (4), http://cercor.oxfordjournals.org/cgi/content/full/bhn080/DC1). We used our previous unsmoothed frontopolar cortex (BA 10) region created using the WFU pickatlas tool implementing the Talairach Daemon atlas (7). To be consistent with the wider literature on MDD (8), we constrained the previous ROI which included AAL hippocampus and parahippocampal regions to the hippocampus only. Thus the following right hemisphere AAL ROIs were used: basal ganglia (i.e. striatum and pallidum), amygdala, insula. The following areas used as ROIs in our previous independent paper (3) were not extracted due to their poor anatomical specificity rendering them useful for small volume correction, but not for extracting average effects: AAL posterior superior temporal sulcus/temporoparietal junction, ventromedial, dorsolateral and dorsomedial prefrontal cortex. The following areas were also not used: the ventral tegmental area, because it is not directly connected with the ATL, and the septal region because its overlap with our smoothed SCC ROI.
